# Application of Mechanical Quantitative Techniques in Postoperative Rehabilitation Assessment of Anterior Cruciate Ligament Reconstruction : a study protocol

**DOI:** 10.1101/2025.05.07.25326783

**Authors:** Zhichun Zhu, Jianyun Zhang, Wenxiang Ouyang, Tiantian Li, Jinsong Xiong, Mi Fan, Jing Guo, Peng Wang, Yongqing Tong, Yulu Yao

## Abstract

This study protocol aims to explore the application of mechanical quantitative techniques in the postoperative rehabilitation assessment following anterior cruciate ligament reconstruction (ACLR). Anterior cruciate ligament injuries are prevalent among athletes and pose a considerable threat to their careers. While ACLR remains the primary therapeutic intervention for such injuries, the assessment of postoperative rehabilitation still encounters significant challenges. Traditional assessment methods are limited by subjectivity and the absence of quantitative data. Mechanical quantitative techniques present a novel approach, offering objective and precise quantitative metrics for knee functional rehabilitation through the analysis of soft tissue mechanical properties. Methods: An observational methodology will be employed to gather clinical data from patients with ACL injuries. A soft tissue mechanical quantitative measurement device will be utilized for knee functional assessment. The anticipated sample size is 66 cases, with inclusion criteria encompassing specific age ranges, injury durations, and other pertinent factors. Exclusion criteria will exclude participants with other injuries or diseases that may impact rehabilitation. The assessment protocol will consist of traditional knee functional assessments, including knee muscle strength, range of motion, and Lysholm scores, as well as mechanical quantitative assessments of the quadriceps and hamstrings. Blinding: Due to the inherent challenges in clinical controlled trials, a strict double-blind design is infeasible. Therefore, participants will be aware of their group assignments, whereas assessors will remain blinded to the specific research objectives and group allocations. Data entry and statistical grouping will be managed by independent personnel. Statistical Analysis: Data analysis is conducted using SPSS 20.0 software. Categorical data are represented as frequencies (percentages), with intergroup comparisons made using chi-square tests or exact probability methods; continuous data are represented as means ± standard deviations, with intergroup comparisons made using t-tests, analysis of variance, or rank sum tests, and correlation analysis is performed using Spearman’s correlation coefficient. A P-value <0.05 is considered statistically significant. Expected Results: Firstly, It is anticipated that there will be a significant correlation between the assessment results of mechanical quantitative techniques and traditional functional assessment methods. This will validate the reliability of mechanical quantitative techniques and facilitate their complementary use alongside traditional assessment methods. Secondly. It is expected that this study will develop new scientific measurement tools and methods, mitigating errors associated with subjective judgments and ensuring the objectivity and accuracy of assessment results. By providing specific, quantifiable data, this study will offer physicians a more intuitive and precise basis for assessment.

## 1. Introduction

### 1.1 Anterior Cruciate Ligament Injury is Becoming an Increasingly Significant Health Issue

Anterior cruciate ligament (ACL) injuries are relatively common sports injuries among athletes, posing a severe threat to their competitive performance and athletic careers [1]. In the United States, for instance, over 350,000 athletes and sports enthusiasts annually experience ACL injuries [2], with approximately half necessitating surgical intervention [3]. The ACL’s physiological attributes impart it with limited self-repair capabilities, leading to the clinical consensus that complete (grade III) ACL tears necessitate anterior cruciate ligament reconstruction (ACLR) [4]. The primary objective of ACLR is to reinstate knee joint stability and postpone the progression of osteoarthritis [5]. Despite significant advancements in surgical techniques for ACL injuries, existing literature indicates a substantial risk of second ACL ruptures post-ACLR, with a prevalence rate as high as 25% among athletes [6]. This elevated risk is primarily attributed to inadequate functional recovery and biomechanical stability post-surgery, rendering the knees vulnerable during high-impact activities [7]. Therefore, more scientific and precise assessment methods are needed post-ACLR to ensure athletes can safely return to sports and reduce the risk of a second ACL rupture.

### 1.2 The Importance of Knee Joint Function Assessment

Knee function assessment plays a pivotal role in the management and rehabilitation of patients with anterior cruciate ligament (ACL) injuries. Accurate evaluation enables clinicians to determine the severity of ACL injuries, providing a critical foundation for the development of personalized treatment strategies. During rehabilitation, routine assessments not only facilitate the monitoring of patient progress and ensure the efficacy of the rehabilitation protocols but also enable the early detection and intervention of potential complications such as muscle atrophy and joint stiffness. Furthermore, periodic evaluation improve patient awareness of their recovery status, fostering greater confidence in treatment, enhancing adherence to rehabilitation programs, and ultimately improving overall quality of life.

Currently, knee function assessment primarily relies on clinical rating scale. While these scales are widely adopted due to their simplicity and ease of administration, their reliability is often compromised by inter-individual variability, assessor subjectivity, and dependence on clinical experience. This limitation is particularly evident in muscle tone assessment, where obtaining objective quantitative data remains challenging [8]. Several studies suggest that conventional spasticity assessment scales fail to accurately reflect joint angular velocity or range of motion [10]. Similarly, manual muscle testing is inherently subjective. Although isokinetic strength testing is considered the “gold standard” for muscle strength quantification, its clinical application is constrained by the requirement for costly, bulky equipment and the logistical challenges associated with patient positioning and transfer [9].

Imaging modalities constitute another essential component of knee function assessment. Traditional imaging techniques such as X-ray and computed tomography (CT) provide high-resolution visualization of bone morphology; however, their two-dimensional nature limits the accurate representation of soft tissue structures [12]. Additionally, these modalities present inherent risks of radiation exposure and may cause pain during the examination process [11]. While magnetic resonance imaging (MRI) offers superior diagnostic accuracy for soft tissue evaluation, its clinical utility is limited by the necessity for multi-sequence, multi-plane scanning, which significantly increases healthcare costs and requires advanced technical expertise. Moreover, MRI is unsuitable for real-time dynamic assessment of functional movement. Given these limitations, there is a compelling clinical need for the development of more precise, real-time, and objective methodologies for comprehensive knee function assessment.

### 1.3 Application of Musculoskeletal Ultrasound in the Assessment of Anterior Cruciate Ligament Injuries

In recent years, musculoskeletal ultrasound has emerged as a focal point in clinical research for knee joint function diagnosis. It is non-invasive, real-time, convenient, and radiation-free [11], offering advantages such as ease of operation and low cost, which have contributed to its widespread clinical application [13]. Musculoskeletal ultrasound provides a clear visualization of the structural characteristics of the knee joint, allowing dynamic observation of muscles and tendons [14]. It enables precise assessment of anatomical abnormalities and functional changes in the knee joint, serving as a valuable reference for personalized rehabilitation treatment [15].

Shear wave elastography (SWE), a recently developed technology, enhances traditional ultrasound capabilities by sensitively detecting changes in tendon hardness. Building upon high-frequency ultrasound, SWE more effectively identifies minor tears [16] and objectively evaluates muscle tissue hardness variations [17]. However, despite its advantages, SWE technology faces several limitations in clinical application. Operating SWE requires experience with two-dimensional ultrasound [18], hindering its broader adoption. Additionally, measurement results can vary across imaging devices from different manufacturers, with studies showing stastistically significant differences in shear wave speed (SWS) [19]. Furthermore, the calculation model for SWE assumes the test subject is a uniform, isotropic elastic body [20], yet musculoskeletal tissue exhibits clear anisotropic properties, failing to meet the conditions of Young’s modulus formula. The order of magnitude changes in Young’s modulus after conversion further increase measurement variability [21]. Some researchers even suggest that SWE data presents extraction and interpretation challenges, with unclear physical significance [22].

### 1.4 Advantages and Research Progress of Mechanical Quantitative Techniques in the Assessment of Anterior Cruciate Ligament Injuries

The propagation characteristics of waves in biological soft tissues under dynamic mechanical loads demonstrate intricate behaviors that are closely linked to the tissues’ mechanical properties. In particular, the velocity of shear wave propagation serves as a key indicator of tissue stiffness: the faster the wave propagates, the stiffer the tissue.[23]. By meticulously analyzing the in vivo propagation patterns of different wave modes, researchers can quantitatively characterize the mechanical properties of soft tissues. This approach enables a non-invasive and non-destructive assessment of both physiological and pathological states of human soft tissues, providing new perspectives for disease diagnosis, prognosis, and broader public health implications. Due to the inherent variability in human tissues properties, precisely characterizing their mechanical behavior remains a major challenge in biomechanics.

Building upon these wave propagation principles, quantitative musculoskeletal biomechanics assessment integrates sophisticated biomechanical modeling with the critical clinical challenge of knee joint function evaluation in patients with ACL injuries. By employing non-destructive quantitative biomechanical measurements, this approach establishes direct quantitative correlation between the degree of knee joint functional recovery and the mechanical state of skeletal muscles in ACL-injuried patients. Consequently, it introduces a novel clinical evaluation framework for knee joint recovery that overcomes the subjectivity of traditional grading scales, thereby providing a more precise and objective assessment. Moreover, the musculoskeletal biomechanical measurement device is user-friendly, non-invasive, and painless, making it suitable for rehabilitation assessments across all age groups. The data and findings from this research are expected to significantly advance rehabilitation protocols and establish evidence-based guidelines for the clinical management of ACL injury patients.

## 2. Materials and methods

### 2.1 Clinical trials

This study primarily employs an observational approach, focusing on musculoskeletal mechanical quantitative detection technology, and includes the following steps: Firstly, collect clinical data from patients with anterior cruciate ligament injuries, including medical history, imaging examinations, etc., to ensure the diversity and representativeness of the sample. Secondly, use musculoskeletal mechanical quantitative measurement devices to conduct quantitative, non-destructive measurements of the patient’s knee joint to obtain relevant parameters of the mechanical state of skeletal muscles. During the measurement process, strictly adhere to operational protocols to ensure the accuracy and reliability of the data. Then, conduct traditional rehabilitation assessments of the knee joint. Concurrently, combine quantitative modeling and characterization methods of biomechanics to deeply analyze the measurement data, revealing the quantitative correlation between the degree of knee joint functional recovery in patients with anterior cruciate ligament injuries and the results of traditional rehabilitation assessments. Finally, based on the analysis results, propose a new clinical evaluation method for knee joint functional recovery and conduct validation and assessment. Additionally, collect subjective feedback from patients to further verify the accuracy and feasibility of the new evaluation method.All trial procedures and radiological and clinical visits are summarized in Figure 1a, b. The research is planned to conduct from 11 March 2025 (first patient included) until 29 October 2025(last follow-up).This research has been registered in ISRCTN (ISRCTN31723656) and OSF (DOI:doi.org/10.17605/OSF.IO/K24QV).And the Ethics Committee of Hunan Provincial Rehabilitation Hospital has approved this research at 11 October 2024 (Approval No.:2024101101).All participants will sign the informed consent in writing, and the signed informed consent will be kept by the research group.

**Figure 1a.**
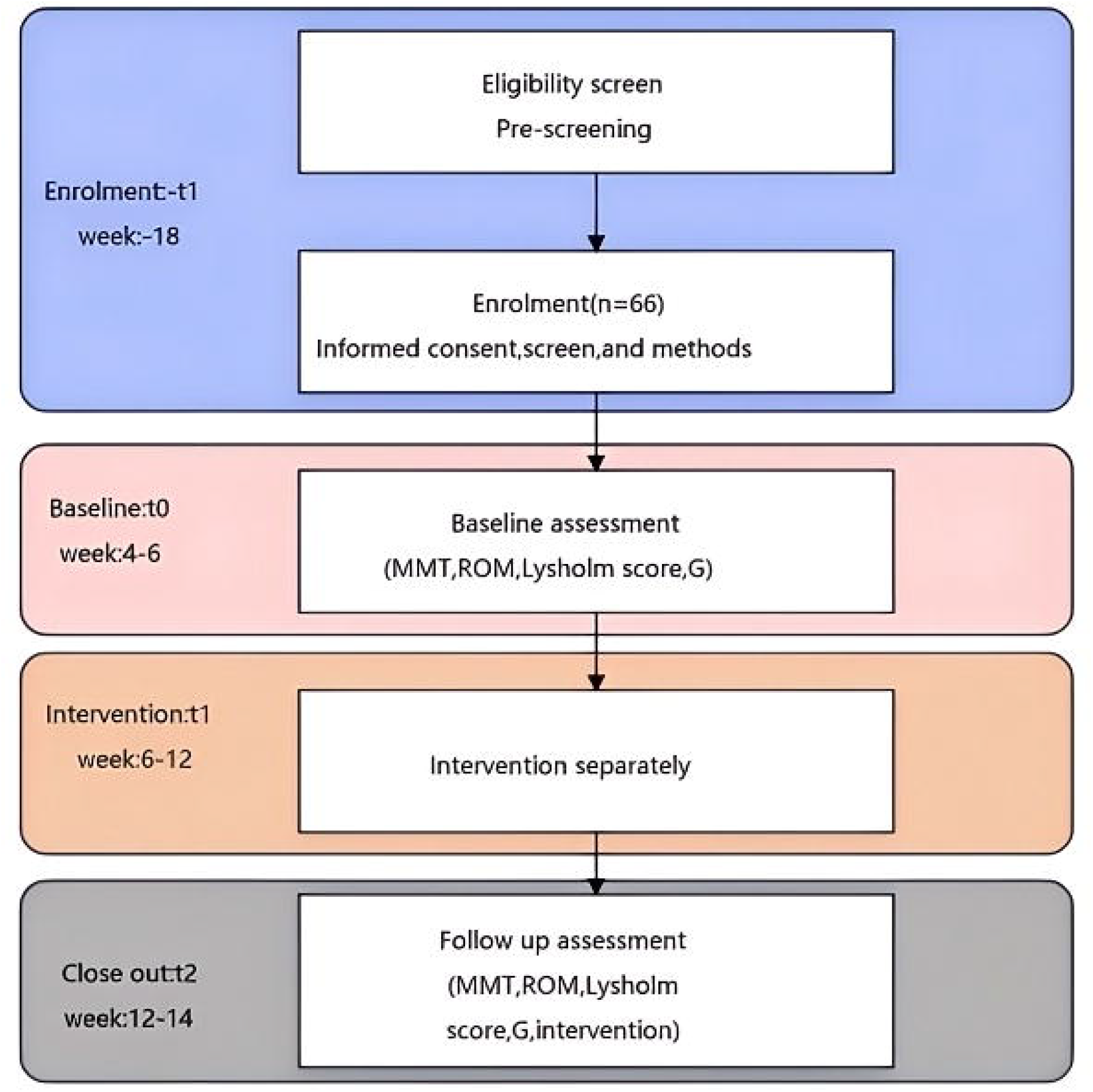
Participant timeline.

**Figure 1b.**
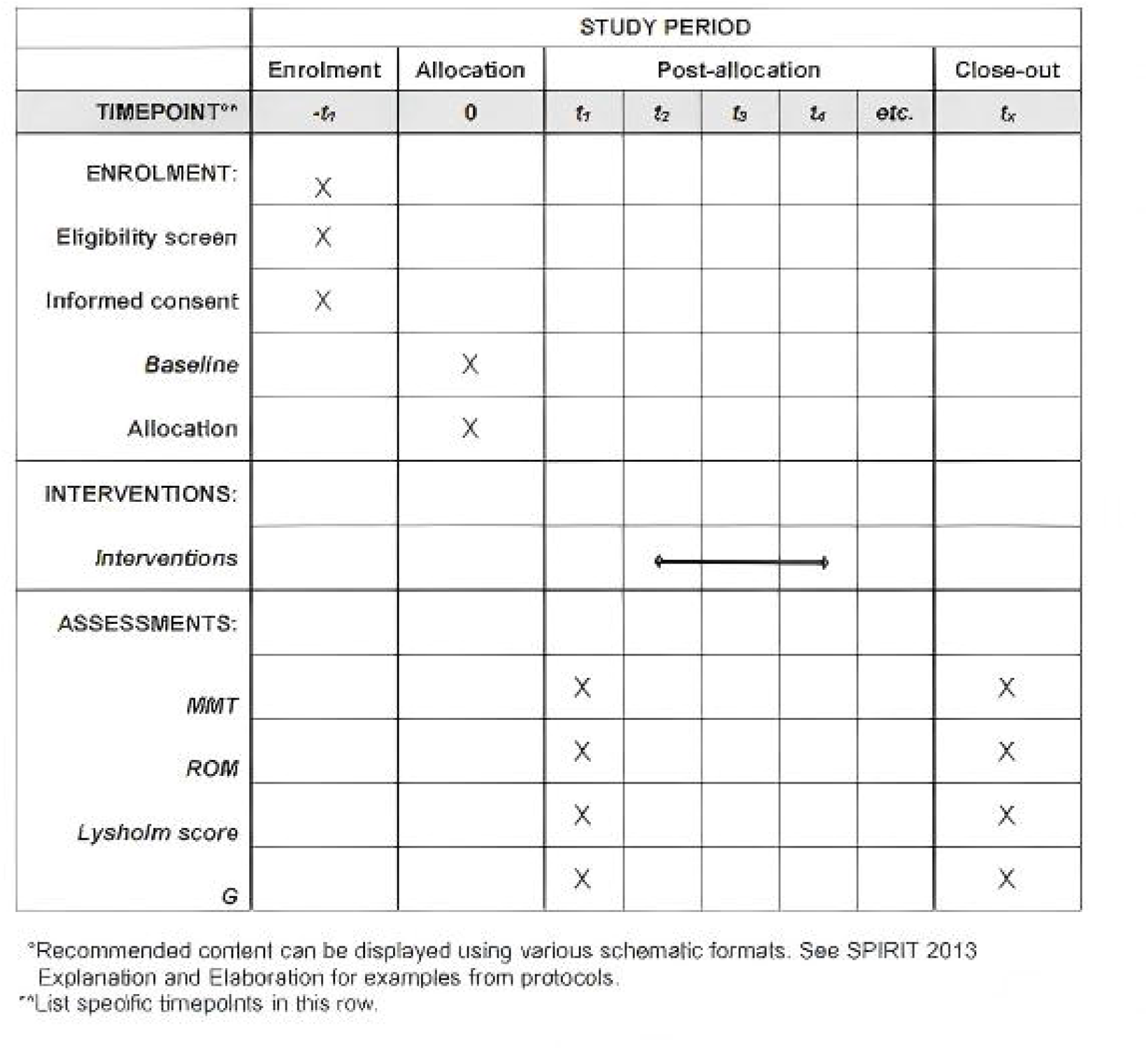
Timing of all procedures.

### 2.2 Experimental study design

#### 2.2.1 Sample recruitment

##### Inclusion criteria

⍰Unilateral leg ACL injury, arthroscopic autologous hamstring or patellar tendon ACL reconstruction surgery, with no other ligamentous injuries involved;

⍰Swelling of the affected knee joint is grade 0 or 1+;

⍰Time from injury to surgery is less than 2 months;

⍰Ages between 18 and 60 years;

⍰Signed an informed consent form for rehabilitation treatment and actively cooperate with the treatment;

⍰The treatment plan is approved by the Ethics Committee of the Hunan Provincial Rehabilitation Hospital;

##### Exclusion criteria

⍰Concurrent posterior cruciate ligament rupture or meniscal repair;

⍰Concurrent postoperative complications affecting limb exercise;

⍰Patients with a history of hip joint trauma;

⍰Individuals with hypertension, diabetes, or other chronic diseases of organs;

⍰Individuals with concurrent severe diseases of the heart, brain, kidneys, and hematopoietic system, and patients with mental illnesses;

⍰Concurrent with any conditions that are detrimental to patient recovery or continuation of the trial.

##### Sample size

In this study, the Lysholm knee score questionnaire comprises a total of 8 variables, basic functional assessment includes 2 variables: muscle strength and range of motion, mechanical quantitative assessment results include 2 variables, a total of 12 research variables are included in this study, referring to Kendall’s sample size estimation method, the sample size included is 5 to 10 times the number of variables, considering a 10% rate of ineligible cases, the total sample size should be at least 66 cases, according to the actual situation, the final sample size is determined to be 66 knee joints.

#### 2.2.2 Implementation method

1. Data Statistics and Analysis:□Firstly, a designated person evaluates the cases that meet the inclusion criteria and records the baseline data;□One assessment is conducted during the rehabilitation process and after treatment, with proper data recording;□Finally, a professional separate from the assessment work performs data statistics and analysis.(2) Knee Joint Function Assessment: A fixed experienced intermediate rehabilitation therapist conducts the functional assessment, which includes: knee joint muscle strength, ROM, and Lysholm score.
2. Knee Joint Muscle Strength Grading: During the examination, the patient is placed in different positions to be tested, and the targeted muscles or muscle groups perform specific movements under conditions of weight reduction, gravity resistance, or resistance, achieving the maximum range of motion. Based on the muscle’s ability to perform the movement, muscle strength is graded according to the grading standards, which are mainly divided into 6 levels: 0, 1, 2, 3, 4, and 5. Level 5 represents normal strength. Lysholm Score:□Pain Score: 0-25 points.□Instability Score: 0-25 points.□Squatting Score: 0-5 points.□Locking Score: 0-15 points.□Climbing Stairs Score: 0-10 points.□Support Score: 0-5 points.□Swelling Score: 0-10 points.□Gait Score: 0-5 points.
3. Musculoskeletal Mechanical Quantitative Assessment: A physician with over 5 years of experience is designated to conduct musculoskeletal mechanical quantitative assessments, The assessment includes musculoskeletal mechanical quantitative testing:□Shear modulus of the rectus femoris muscle (modulus of rigidity, G);□Shear modulus of the hamstring muscles (modulus of rigidity, G). Specific operation method:

a. Measurement Method: Utilize a musculoskeletal mechanical quantitative detector (M5) to measure the mechanical quantitative characteristics (shear modulus G) of the subject’s rectus femoris and hamstring muscles.
b. Subject Position: The patient is in a supine position with both lower limbs relaxed.
c. Operating SOP(Standard Operating Procedure):

Protocol: Experimental Equipment, Consumables, and Paper Documents:

□Experimental Equipment: Musculoskeletal mechanical quantitative detector, including the main unit, 9L3-8.5MHZ linear array transducer, and mechanical excitation module.

⍰Consumables: Ultrasound coupling gel.

⍰Paper Documents: Informed consent form for subjects, subject information collection form, and experimental record form.

⍰Other Experimental Tools and Consumables (including a standard medical bed, measuring tape, power strip, ultrasound-specific paper, marker, heat patch, storage box, low stool, etc.).

##### Pre-experimental Preparation

⍰Coupling Gel Preheating: One hour before the official start of the experiment, attach a heat patch to the coupling gel bottle to warm it to a temperature similar to body temperature for the formal experiment;

⍰Paper Document Preparation: Print paper documents according to the number of subjects, one per person;

⍰Subject Informed Consent: The experimenter informs the subjects about the experimental process, benefits, risks, confidentiality plan, etc., and after being fully informed, the subjects sign the informed consent form;

⍰Subject Completes Information Collection Form; Instrument Preparation

⍰Place the musculoskeletal mechanical quantitative detector and its accessories on the experimental table, ensuring the main unit is properly connected to the transducer; place the power strip in the experiment box, connect the main unit to the power strip. Press and hold the power button on the side of the musculoskeletal mechanical quantitative detector until the power indicator light turns on, open the app, and confirm login.

⍰Tap the E button on the left sidebar of the main unit screen to enter the force-sound acquisition mode, and check the parameters as shown in the table 1 below:

**Table 1.** Parameters.

| Parameter | Value |
| --- | --- |
| Frequency | 6.0M |
| Line Count | 4 |
| Range | 5mm |
| Time | 300ms |
| Position Preset | 2 |
| Display Line Number | Off |

⍰Adjust the depth position of the measurement point caliper to set the starting point of the acquisition depth range just below the dermis layer under the transducer as shown in the figure 2:

**Figure 2.**
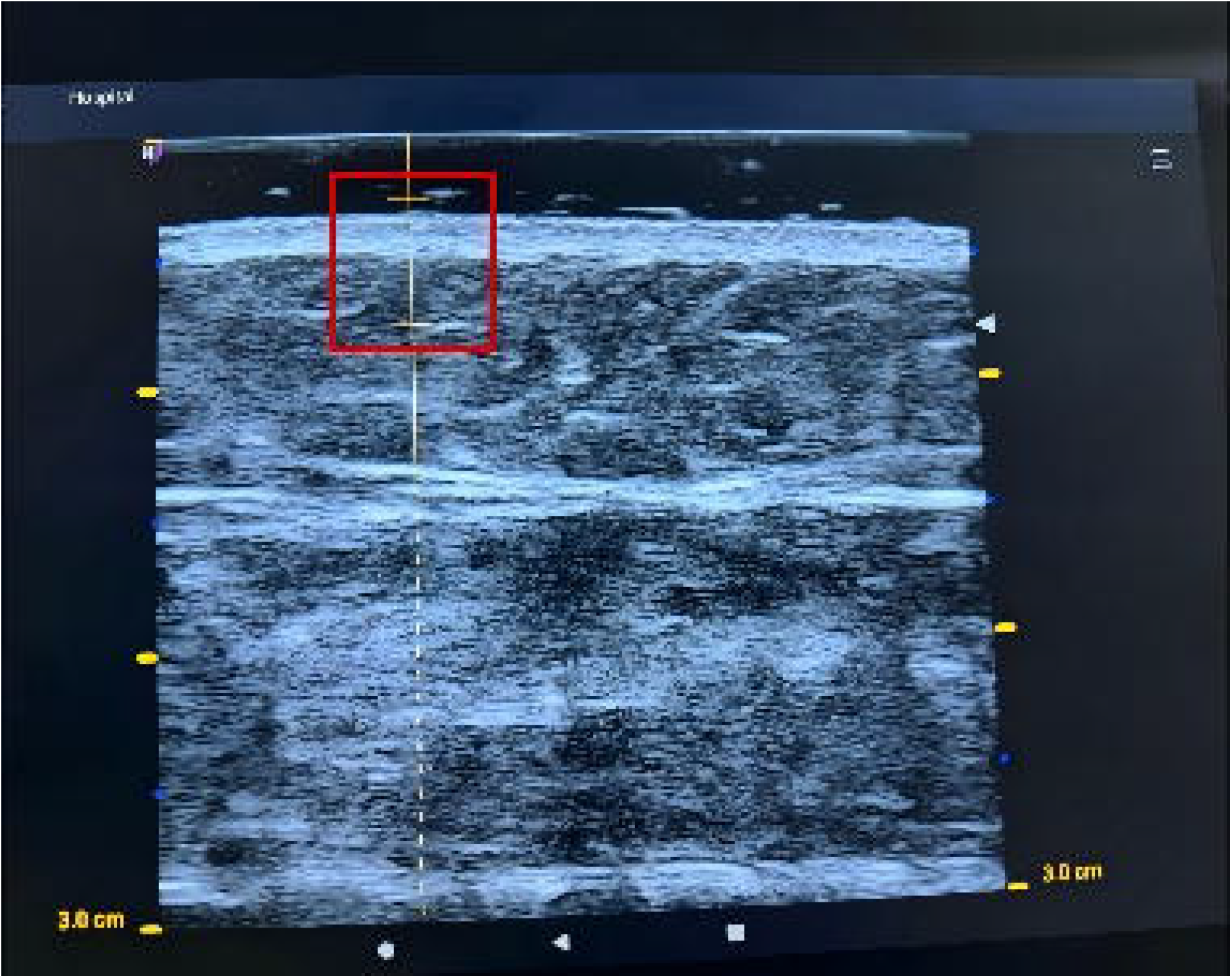
starting point of the acquisition depth range.

⍰After the settings are completed, click on the B button on the left sidebar of the main unit screen to enter B-mode ultrasound, and complete the preparation work. Experimental Procedure

⍰Guide the subject to prepare for the test, evenly apply the pre-warmed coupling gel on the ultrasound transducer of the musculoskeletal mechanical quantitative detector, place the ultrasound transducer close to the measurement site so that the measurement point is directly below the front half of the transducer (the front is marked by a protrusion on one side of the ultrasound transducer), and observe the imaging effect in B-mode.

⍰Under clear B-ultrasound imaging conditions, use the ruler tool to measure and record the thickness from the skin surface to the corresponding muscle fascia layer to be tested.

⍰Turn on the excitation device switch, gently place the excitation end 3-6mm directly in front of the protrusion side of the transducer, ensuring that the excitation end is perpendicular to the imaging plane of the ultrasound transducer, as shown in the figure 3 below.

**Figure 3.**
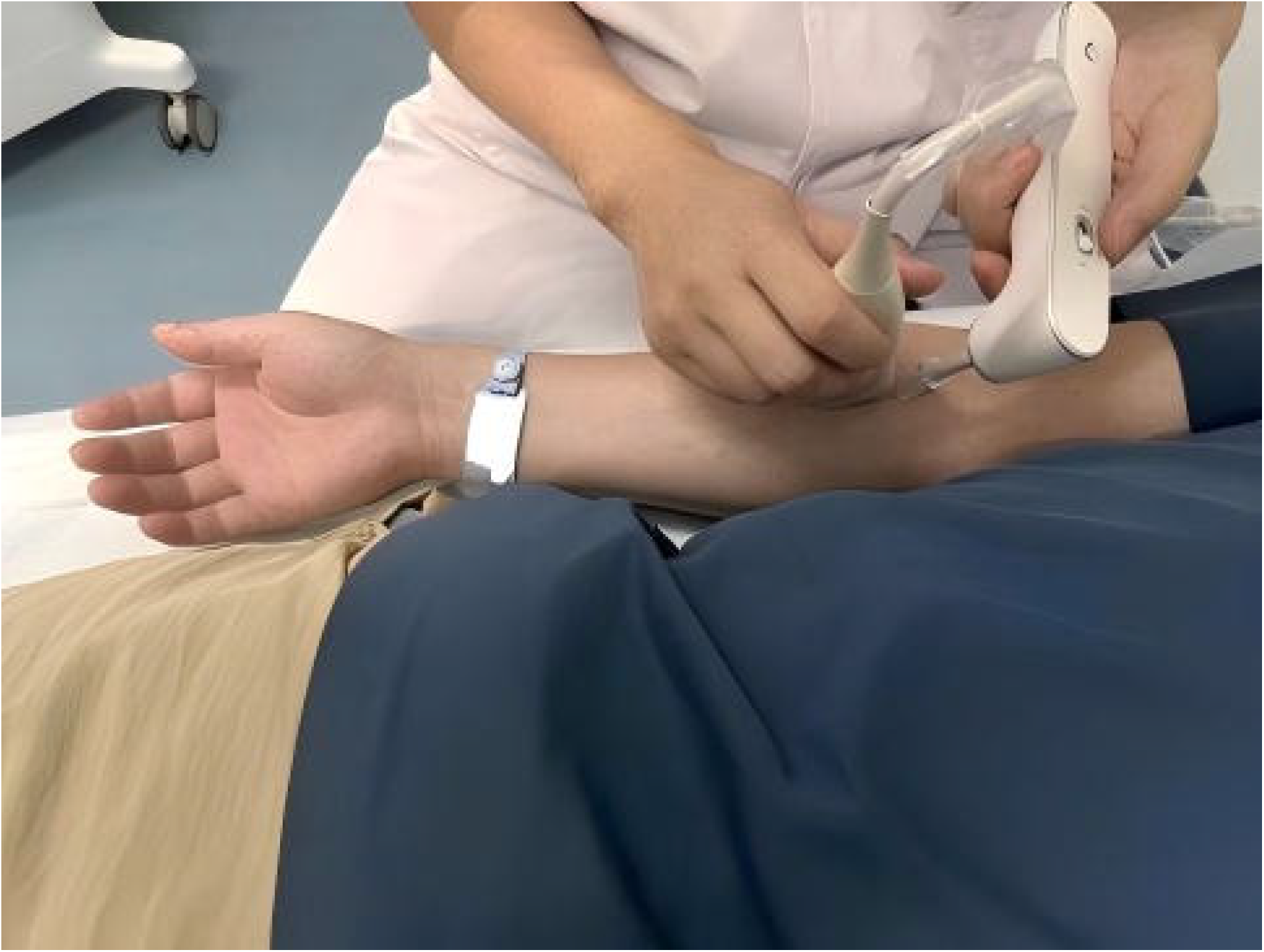
Operation demonstration.

⍰Switch the musculoskeletal mechanical quantitative detector to E-mode to begin the measurement; the valid measurement values will be displayed on the screen interface.

⍰Record the measurement values in the corresponding sections of the experimental record form, completing this measurement experiment.

d. Standards for Data Collection:

⍰Locate the measurement point: Mark the measurement point on the muscle surface according to the study design to ensure consistency in measurement location each time.

⍰Apply coupling gel: Uniformly apply coupling gel to the measurement area to reduce signal attenuation.

⍰Each measurement should obtain no fewer than 5 valid measurement values, and the standard deviation of the measurement values should be less than 10% of the mean measurement value.

#### 2.2.3 Grouping scheme

In a case-control design, the traditional rehabilitation assessments implemented for the knee joint include: 1. Knee joint muscle strength; 2. Range of motion (ROM) of the knee joint; 3. Lysholm score. The mechanical quantitative assessment team conducts mechanical quantitative evaluations on the knee joint.

#### 2.2.4 Blind method

Due to the particularities of clinical controlled trials, a strict double-blind trial cannot be conducted. Therefore, based on the characteristics of this study, participants are aware of their group assignments, but evaluators are not informed of the specific research objectives and group allocation scheme. Data entry and statistical grouping personnel are also set up independently.

#### 2.2.5 Statistical analysis scheme

The statistical analysis plan involves comparing the knee joint function assessment and mechanical quantitative modulus values, as well as the assessment results between the two sides. A statistical analysis software, SPSS 20.0, is used to establish a database. Categorical data are represented as frequencies (percentages), and group comparisons are made using chi-square tests or exact probability methods. Continuous data are represented as means ± standard deviations, and group comparisons are made using t-tests, analysis of variance, or rank sum tests. Correlation analysis of related factors is performed using Spearman’s correlation coefficient. A P-value< 0.05 is considered statistically significant.

#### 2.2.6 Assessment tool description

Mechanical Quantitative Measurement Device (M5) Manual Muscle Testing (MTT Muscle Strength Chart) Range of Motion Assessment (using a Goniometer) Knee Joint Function Assessment Scale (Lysholm Scale)

#### 2.2.7 Expected results

Mechanical quantitative technology, with its unique advantages, has shown extraordinary potential and value in the assessment of knee joint function. This technology not only provides objective and precise quantitative data but also deeply reveals the intrinsic connections between its assessment results and traditional functional assessment methods, making it an indispensable and effective tool for evaluating knee joint function. We anticipate that this study will contribute to knee joint function assessment by 1. discovering a significant correlation between the assessment results of mechanical quantitative technology and traditional functional assessment methods. By verifying the reliability of mechanical quantitative technology through correlation, it also provides the possibility for the complementary application of both assessment methods.2. forming new scientific measurement tools and methods, avoiding errors brought by subjective judgments, ensuring the objectivity and accuracy of assessment results, and providing concrete, quantifiable data, which offers doctors a more intuitive and accurate basis for assessment.

In summary, mechanical quantitative technology has demonstrated tremendous potential and value in the assessment of knee joint function due to its unique advantages. We have reason to believe that in the future, this technology will bring blessings to more patients and contribute significantly to the advancement of medical science.

## Data Availability

All data generated in the future of this study can be provided at the reasonable request of the author

